# Healthcare utilization and cost of care for a multidisciplinary integrated practice unit model of spine rehabilitation compared to standard or no physical therapy

**DOI:** 10.1101/2025.06.10.25327236

**Authors:** Bahar Shahidi, Connor Richards, Lissa Taitano, Armin Zavareh, Kamshad Raiszadeh

**Affiliations:** UC San Diego Department of Orthopaedic Surgery, La Jolla CA USA; Livara Health, San Diego CA USA

**Keywords:** cost effectiveness, spine pain, low back pain, neck pain, physical therapy, economic evaluation

## Abstract

**Objective:** To compare healthcare utilization and cost in individuals with spine pain who undergo no physical therapy, standard physical therapy, or physical therapy in an integrated practice unit model.

**Design:** A cost-effectiveness analysis

**Setting:** Multi-site outpatient physical therapy clinics within a single metropolitan region

**Participants:** Individuals with de-identified claims data from a single insurance provider under Medicare Advantage with a spine-pain related diagnosis from January 2019-December 2021.

**Interventions:** Patients were categorized into three cohorts: No physical therapy (NoPT), standard physical therapy (SPT), and physical therapy within an integrated practice unit model (IPUPT) based on their physical therapy history during the data collection period.

**Main Outcome Measures:** Number and percentage of patients reporting claims, number of claims/patient per year, paid amount, and number of RVUs were compared across groups using chi-square or one-way ANOVA with multiple comparisons corrections.

**Results:** Data from 13,569 patients was included in this study. The number of patients with spine-related inpatient claims was highest in the SPT group (2.8%) compared to the IPUPT (1.5%) and NoPT (1.3%) groups (p=0.004). Outpatient care utilization was driven by radiology (54.7%) and laboratory (22.1%) claims and was lowest in the IPUPT group (N=1,096; 56.8%) compared to the SPT group (N=1,654; 68.3%) and NoPT group (N=9,150; 99.3%, p<0.001). The SPT group was most costly per person ($2,243.66(11,048.94)) followed by the NoPT ($1,352.01(6,419.2), p<0.001) and the IPUPT ($1,259.88(9,061.23), p<0.001) groups. The greatest contributor to cost was outpatient procedures, averaging $142.39(1,046.26) per person.

**Conclusion:** An integrated multidisciplinary rehabilitation model may be a cost-effective method of multimodal care in individuals with spine pain.

## Introduction

Spine pain is one of the most prevalent and disabling musculoskeletal complaints in the general population, putting increasing pressure on the healthcare system for optimizing its management^1–3^. Rising healthcare costs for individuals with spine pain have become a significant concern for health care systems and payers. Escalating healthcare expenditures and reduced reimbursements are putting a strain on healthcare systems and affecting overall population health. With an increasing emphasis on optimizing healthcare outcomes and controlling costs, it is imperative to explore the potential cost savings associated with the most common interventions utilized for treatment of spine pain.

Physical therapy is often one of the recommended treatments for individuals with low back (LBP) or neck pain (NP). Numerous studies suggest physical therapy results in reduced costs in the first year after initial incidence of LBP or NP compared to other interventions^4–7^. Early referral to guideline adherent physical therapy has been associated with 60% lower total LBP-related costs^8^. Additionally, adherence to physical therapy has been associated with improved clinical outcomes and decreased subsequent use of prescription medication, a 50% decrease in medical imaging utilization, and almost 60% decrease in epidural injections^8,9^.

However, these cost savings are thought to be offset by the potential for healthcare overutilization in a condition where a large proportion of individuals are thought to recover within 12 weeks with minimal to no intervention^10^. As a result, clinical guidelines for treatment of spine pain often don’t include a comprehensive physical therapy recommendation until the symptoms become chronic^11,12^. In these cases, the efficacy of rehabilitation is complicated by the multifactorial contributions to development and maintenance of spine pain, including psychosocial factors, occupational and environmental exposures, and underlying comorbidities^13-^15.

To address the complexity of these conditions, many current guidelines have adopted recommendations for interdisciplinary, multimodal, and coordinated care. These approaches include multidisciplinary integrated practice units (IPUs) that incorporate care between surgeons, physical therapists, behavioral specialists, and other wellness experts, and have been shown to effectively reduce pain and disability in these populations^16,17^. However, in practice, these models are less utilized due to the perception that they are resource intensive^18^, and it is unknown whether this model ultimately reduces downstream health care utilization and cost compared to more streamlined conservative care models. The purpose of this study was to compare health care utilization and cost of care across patients with spine pain who receive A) no physical therapy (NoPT), B) standard physical therapy (SPT) or C) physical therapy within an IPU care model (IPUPT).

## Methods

### Participants

De-identified claims data from a single insurance provider (Providence Health and Services) serving 3 constituent medical groups and 4 regional hospital sites was reviewed for individuals receiving health care services associated with a spine-pain related diagnosis. This study was not considered human subjects research due to the de-identified nature of the data and was exempt from Institutional Review Board approval. Individuals with an International Classification of Disease-10^th^ revision (ICD-10) based spine code for health-care claims from January 2019-December 2021 were included. All individuals initiated services under Medicare Advantage under a single insurance provider serving a large metropolitan region from January 2019-December 2020, and data was collected up to December 2021 to generate a minimum of 1-year follow up health care utilization data.

### Intervention groups

Patients were categorized into three cohorts based on their physical therapy history during the data collection period. Individuals who never received physical therapy at any point from January 2019 to December 2021 were assigned to the NoPT group. Individuals who had claims associated with physical therapy within a known IPU-based care network of 5 regional clinics were assigned to the IPUPT group. Individuals who had physical therapy claims associated with community or hospital-based outpatient clinics in the same region were included in the SPT group. Individuals who received physical therapy through both SPT and IPUPT models within the data collection time frame were excluded.

### Cost and claims data

Up to 3 years of health care utilization data was collected on all participants, including spine-and non-spine related inpatient claims, and spine related outpatient claims. All cost and claims data representing health care utilization for the SPT and IPUPT cohorts were analyzed based on the follow up period after completion of the physical therapy program. Among patients meeting the inclusion criteria, spinal, other musculoskeletal (MSK), and non-MSK claims were all considered. Inpatient claims were identified by the diagnosis-related group (DRG) code associated with the claim, and inpatient spinal claims were identified as the surgical DRG codes 28–30, 52, 53, 453–460, 471–473, 518–520, and the non-surgical DRG code 552.

In addition to the cost paid, each outpatient claim was assigned a number of relative value units (RVUs) based on the procedure code, the facility type, and the year in which the claim occurred. The Center for Medicare & Medicaid Services (CMS) Physician Fee Schedule for 2019, 2020, and 2021 was used to assign a relative value unit (RVU) value to each outpatient claim according to these fields^19^. For inpatient claims, the CMS Inpatient Prospective Payment System (IPPS) Final Rule for 2019, 2020^20^, and 2021^21^ was used to assign a number of RVUs to each claim.

Table 5 of the IPPS Final Rule provides a weight factor for each fiscal year. To associate an RVU value, the $35 rate per RVU for current procedural terminology (CPT) claims and $6,122 base payment per DRG were used to assume a conversion rate of 1.00 DRG = 175 RVU. In this way, an RVU value was assigned for each inpatient claim as the weighted DRG value found in the IPPS Final Rule scaled by this 175-conversion factor.

### Statistical Analysis

Number and percentage of patients reporting claims, number of claims/patient per year calculated over the duration of available follow up, paid amount, and number of RVUs were compared across groups using one-way Analysis of Variance with post-hoc Sidak corrections for multiple comparisons for continuous variables, or chi-squared statistics for proportional comparisons. All statistics were performed using SPSS (version 28.0.1.1(14), IBM Corporation, 2021).

## Results

### Participant characteristics

A total of 13,569 patients met the inclusion criteria. The majority of individuals did not undergo physical therapy and were in the NoPT group (N=9,215; 67.9%), followed by SPT (N=2,423; 17.9%), and IPUPT (N=1,931; 14.2%). There was a significantly larger proportion of participants who initiated care in 2019 (N=9,276; 71.7%) as compared to 2020 (N=3,843; 28.3%, p=0.017), which was primarily driven by a lower care initiation rate in the IPUPT setting in 2020 compared to the NoPT group (p<0.006). There were no differences in initiation rates between the IPUPT and SPT groups from 2019 to 2020 (p=0.190). The average follow-up duration for the full cohort was 575.4 (189.7) days, with the NoPT and SPT groups averaging one month (29.4 and 23.8 days respectively) of additional follow up data compared to the IPUPT group (p<0.001).

### Inpatient Claims data

The number of patients with spine-pain related inpatient claims was 220 (1.6%). This was highest in the SPT group (2.8%) compared to the IPUPT (1.5%) and NoPT (1.3%) groups (p=0.004). Similarly, the average (standard deviation) number of spine-related inpatient claims per person was highest in the SPT group (0.02(0.15) claims/year) compared to the IPUPT group (0.01(0.10) claims/year, p<0.001)) and the NoPT group (0.01(0.09) claims/year, p=0.001)), with no significant differences between the NoPT and IPUPT groups (p=0.943).

The number of patients with non-spine related inpatient claims was 96 (0.07%). The number of non-spine related inpatient claims was highest in the NoPT group (N=76; 0.08%), followed by the SPT group (N=14; 0.05%), and the IPUPT group (N=6, 0.03%, p=0.035). The average number of non-spine related inpatient claims per person was not significantly different across treatment groups, although there was a trend for a lower number of claims in the IPUPT group as compared to the NoPT group (p=0.063).

### Outpatient Claims data

The number of patients with non-PT spine-related outpatient claims was 11,900 (87.7%). Outpatient care utilization was lowest in the IPUPT group (N=1,096; 56.8%) compared to the SPT group (N=1,654; 68.3%) and NoPT group (N=9,150; 99.3%, p<0.001). The average number of outpatient claims/person per year was significantly greater in the SPT group (15.2(42.4) claims/year) compared to the IPUPT (9.5(28.0)) and NoPT (11.7(32.7)) groups (p<0.001; **TABLE 1**). The NoPT group also demonstrated significantly higher outpatient claims rates compared to the IPUPT group (p=0.028, **FIGURE 1A-C**).

**TABLE 1.** Number of individuals with claims across claim type and group.

| Group | Spine Inpatient<br>Claims, N(%) | Non-Spine Inpatient<br>Claims, N(%) | Outpatient Claims<br>N(%) |
| --- | --- | --- | --- |
| NoPT (N=9,215) | 123 (1.3%) | 76 (0.08%) | 9150 (99.3%) |
| SPT (N=2,423) | 69(2.8%) | 14 (0.05%) | 1654 (68.3%) |
| IPUPT (N=1,931) | 28 (1.5%) | 6 (0.03%) | 1096 (56.8%) |

**FIGURE 1.**
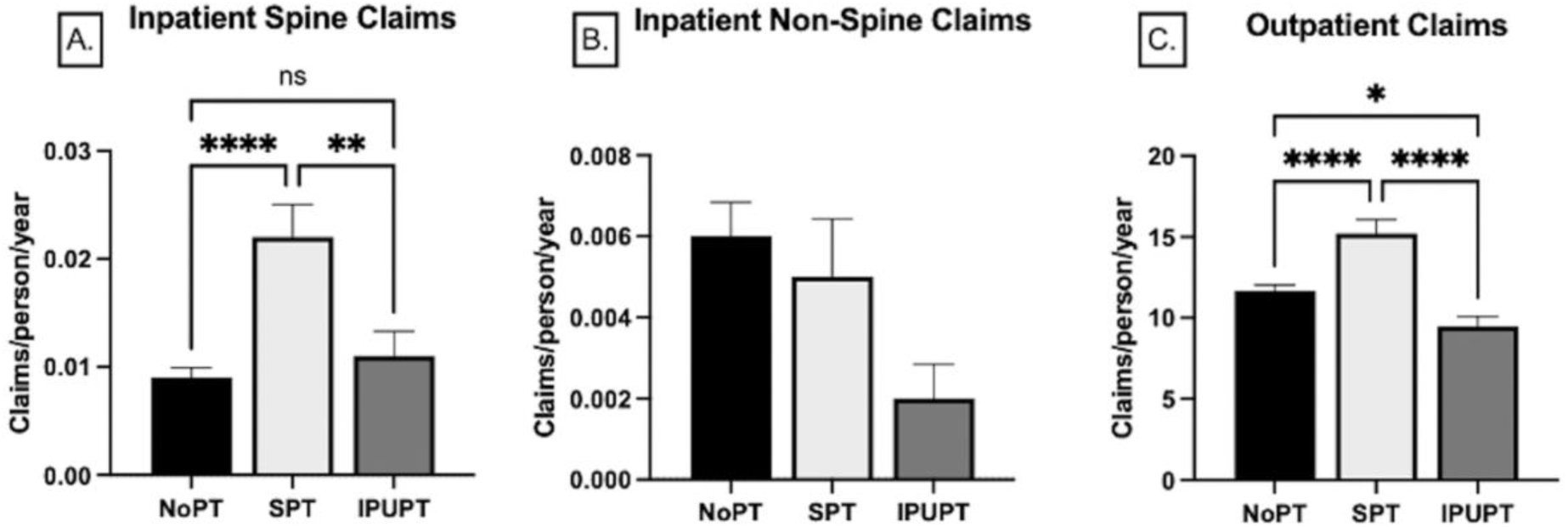
Inpatient Spine(A), Inpatient Non-spine(B), and Outpatient(C) claims/person/year across the NoPT, SPT and IPUPT groups. Data are represented as mean(SE). *(p<0.05), **(p<0.01), ****(p<0.001).

Outpatient claims were most prevalent across all groups in the category of radiology (54.7%), followed by laboratory (22.1%) claims. The least prevalent claims were for chiropractic or osteopathic services (0.04%). The NoPT group had the highest utilization of radiology and laboratory related services, whereas the SPT group had the highest utilization of anesthesiology, chiropractic/osteopathic, outpatient surgery, and outpatient procedures (**TABLE 2**).

**TABLE 2.**
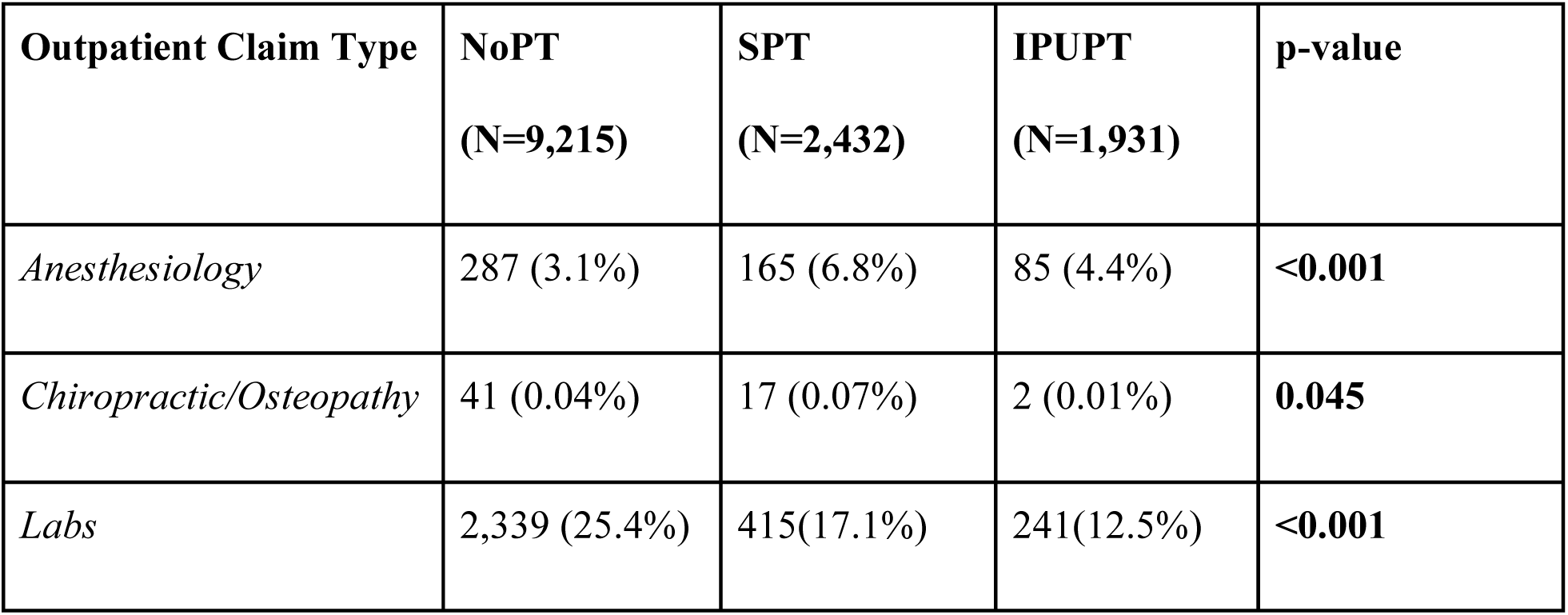
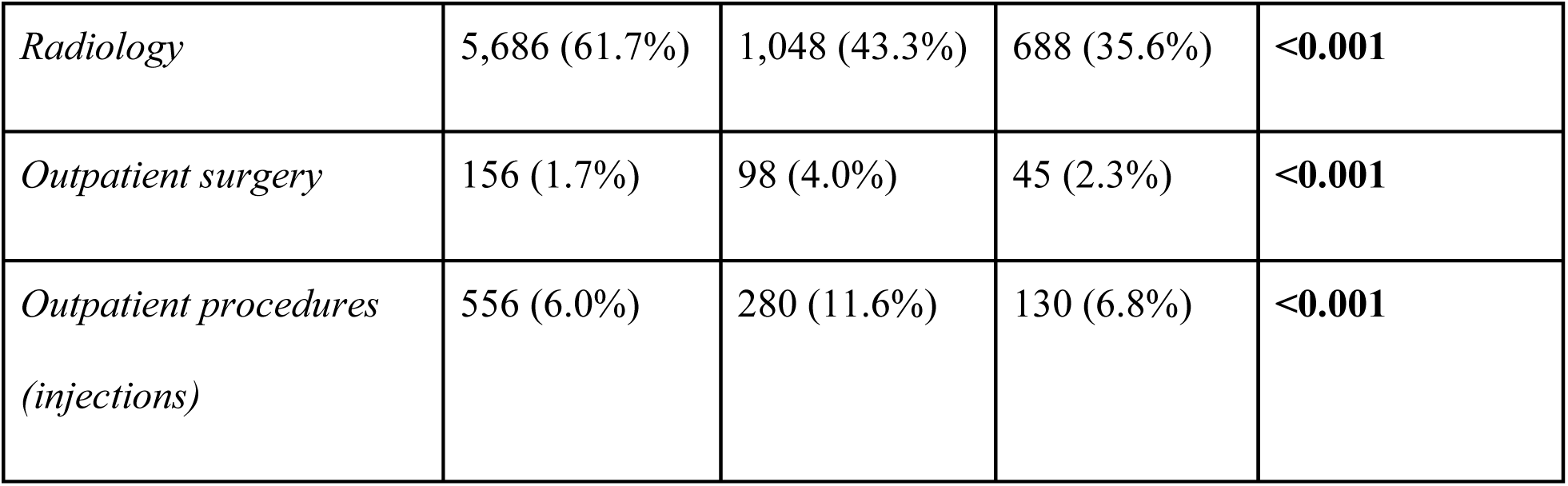
Outpatient claim types according to group. Bold p-values indicate a statistically significant difference in claim prevalence across groups.

| Outpatient Claim Type | NoPT<br>(N=9,215) | SPT<br>(N=2,432) | IPUPT<br>(N=1,931) | p-value |
| --- | --- | --- | --- | --- |
| *Anesthesiology* | 287 (3.1%) | 165 (6.8%) | 85 (4.4%) | **<0.001** |
| *Chiropractic/Osteopathy* | 41 (0.04%) | 17 (0.07%) | 2 (0.01%) | **0.045** |
| *Labs* | 2,339 (25.4%) | 415(17.1%) | 241(12.5%) | **<0.001** |
| *Radiology* | 5,686 (61.7%) | 1,048 (43.3%) | 688 (35.6%) | **<0.001** |
| *Outpatient surgery* | 156 (1.7%) | 98 (4.0%) | 45 (2.3%) | **<0.001** |
| *Outpatient procedures*<br>*(injections)* | 556 (6.0%) | 280 (11.6%) | 130 (6.8%) | **<0.001** |

### Cost of Care

Total cost of care per year across the duration of the study period for all claims (inpatient and outpatient combined) was $20.3 million with an average cost per person of $1,498.12. The NoPT group was responsible for the greatest total cost at $12.5 million (57.3%), followed by the SPT group at $5.4 million (25.0%) and the IPUPT group at $2.4 million (11.2%). When averaged on a per person basis, the SPT group had the highest cost per person ($2,243.66(11,048.94)) followed by the NoPT ($1,352.01(6,419.2), p<0.001) and the IPUPT ($1,259.88(9,061.23), p<0.001) groups. There were no significant differences between the average cost per patient for the NoPT and IPUPT groups (p=0.953, FIGURE 2A,B).

**FIGURE 2.**
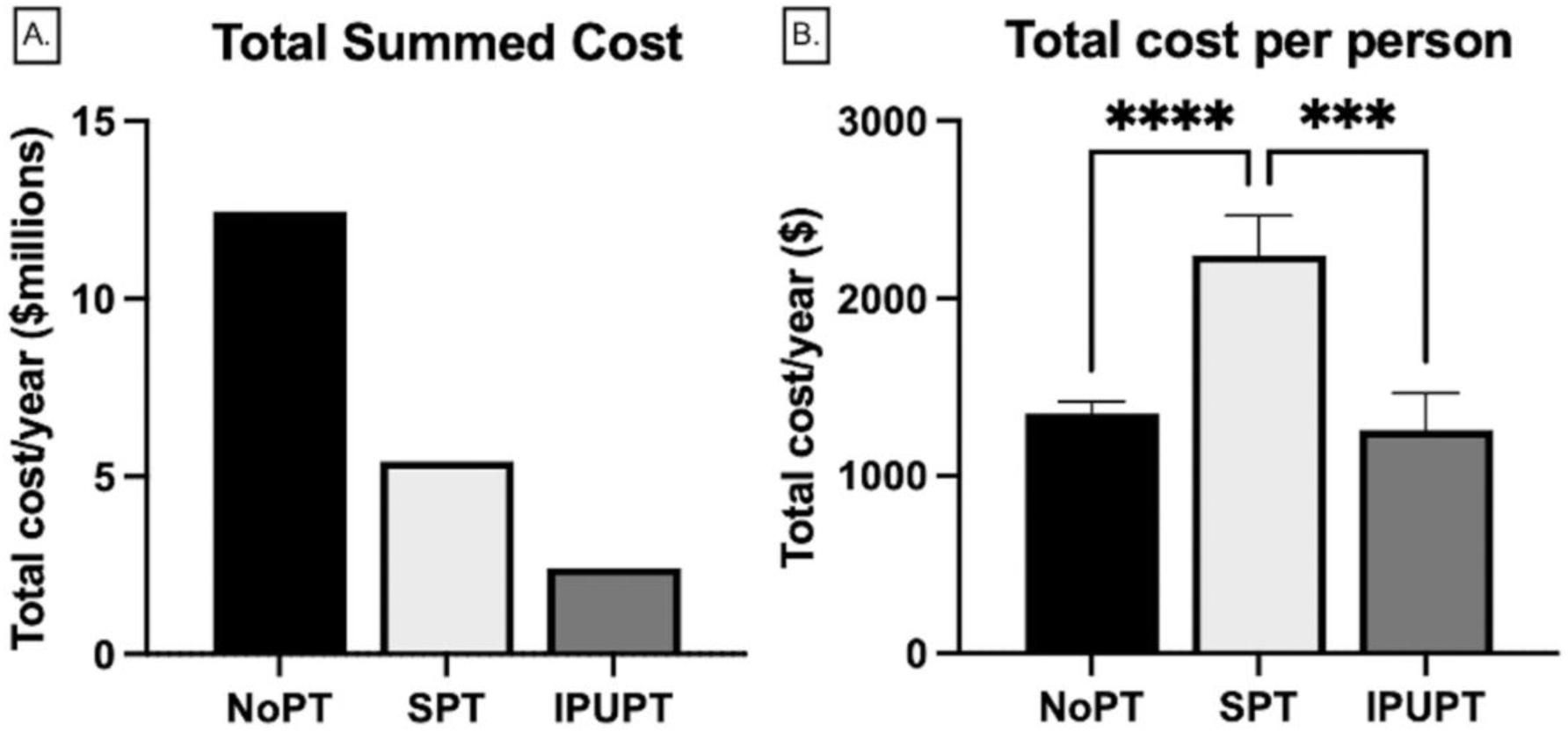
(A) Total cost of care per year across all group participants in millions of dollars. (B) Average annual cost of care on a per/person basis across groups. Bars are mean (standard deviation). ***p<0.001, ****p<0.0001.

Similar overall patterns were observed when costs were split out into in-patient spine related, inpatient non-spine related, and outpatient costs. For inpatient spine-related costs, the NoPT group accounted for the greatest total costs ($3.2 million) followed by the SPT group ($2.3 million) and the IPUPT group ($890,371). On a per person level, the NoPT group had the lowest annual cost ($349.67(4,257.07) compared to the SPT group ($934.50(8,006.50), p<0.001). The SPT group had significantly greater cost compared to the IPUPT group (p=0.017), but there were no differences between the NoPT and IPUPT ($461.09(7,342.88) groups(p=0.812; FIGURE 3A). RVU number was also highest in the SPT group (16.2(111.9)) compared to the IPUPT (7.5(80.0); p<0.001) and NoPT (6.1(64.0); p<0.001) groups. Annual spine-related inpatient cost savings for the IPUPT group compared to the SPT group was $473.41 per person. Spine-related inpatient cost savings for NoPT compared to SPT was $584.83 per person and $111.42 per person compared to IPUPT.

**FIGURE 3.**
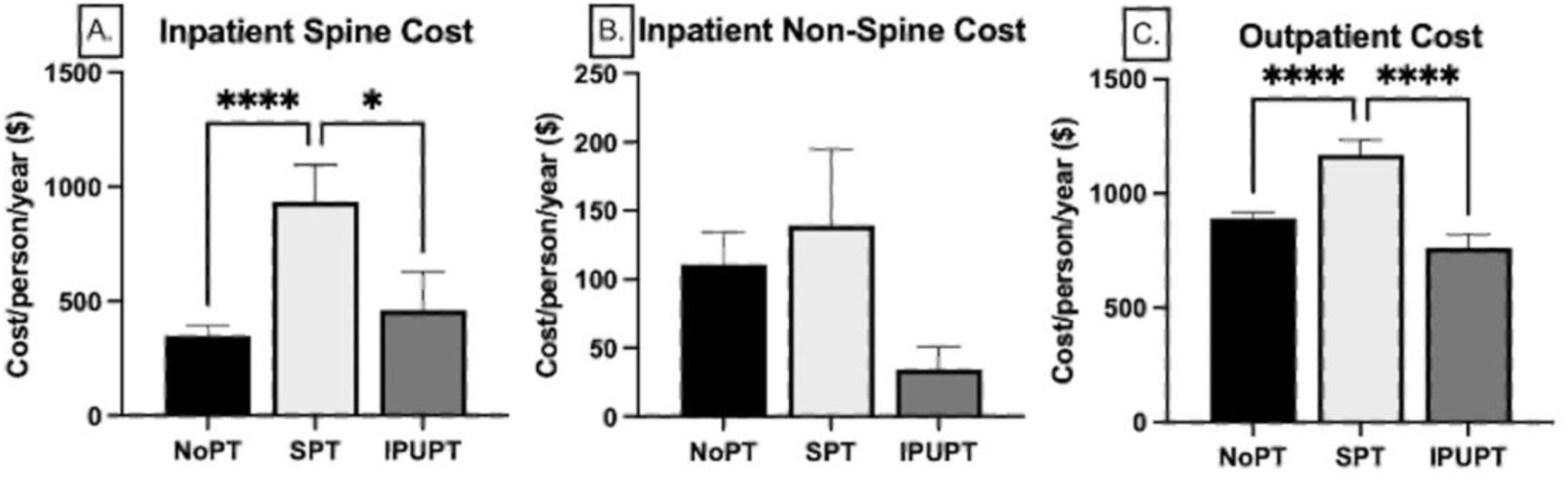
Inpatient Spine(A), Inpatient Non-spine(B), and Outpatient(C) annual cost per person across the NoPT, SPT and IPUPT groups. Data are represented as mean(SE). *(p<0.05), ****(p<0.001).

The total cost for inpatient non-spine related services was $1.4 million and there were no differences in annual costs (p=0.272), or number of RVU’s (p=0.314) per person across groups (FIGURE 3B). Outpatient services drove the majority of healthcare utilization costs for all groups, with total outpatient expenditures amounting to $12.5 million. Similar to inpatient costs, the greatest contributor to outpatient cost was the NoPT group ($8.2 million) followed by the SPT group ($2.8 million) and the IPUPT group ($1.5 million). On an annual per person basis, the SPT group had the highest utilization ($1,169.90(3,156.37) compared to the NoPT ($891.50(2,454.92), p<0.001) and IPUPT ($764.46(2,521.34), p<0.001) groups. There were no differences between the NoPT and IPUPT groups (p=0.146, FIGURE 3C). Outpatient RVU numbers were highest in the SPT group (30.8(100.4)) compared to the IPUPT (16.8(65.6); p<0.001) and NoPT (18.1(59.4); p<0.001) groups. Annual outpatient cost savings for IPUPT compared to SPT was $405.46 per person, and $127.03 per person compared to NoPT. Outpatient cost savings for NoPT compared to SPT was $278.42

Across the outpatient claim categories, the greatest annual expenditures on a per person basis were in outpatient procedures, averaging $142.39(1,046.26) per person, followed by radiology, averaging $122.39(302. 64) per person, whereas chiropractic and osteopathic services were the least expensive at $0.39 (11.82) per person. The NoPT group generated the greatest costs in laboratory, radiology, and chiropractic/osteopathy services, with the lowest costs in anesthesiology, outpatient surgery, and outpatient procedures. The SPT group generated the greatest costs in anesthesiology, outpatient surgery, and outpatient procedure claim categories, and were least expensive in chiropractic/osteopathy and laboratory services. The IPUPT group was the least expensive in radiology (**TABLE 3**).

**TABLE 3.**
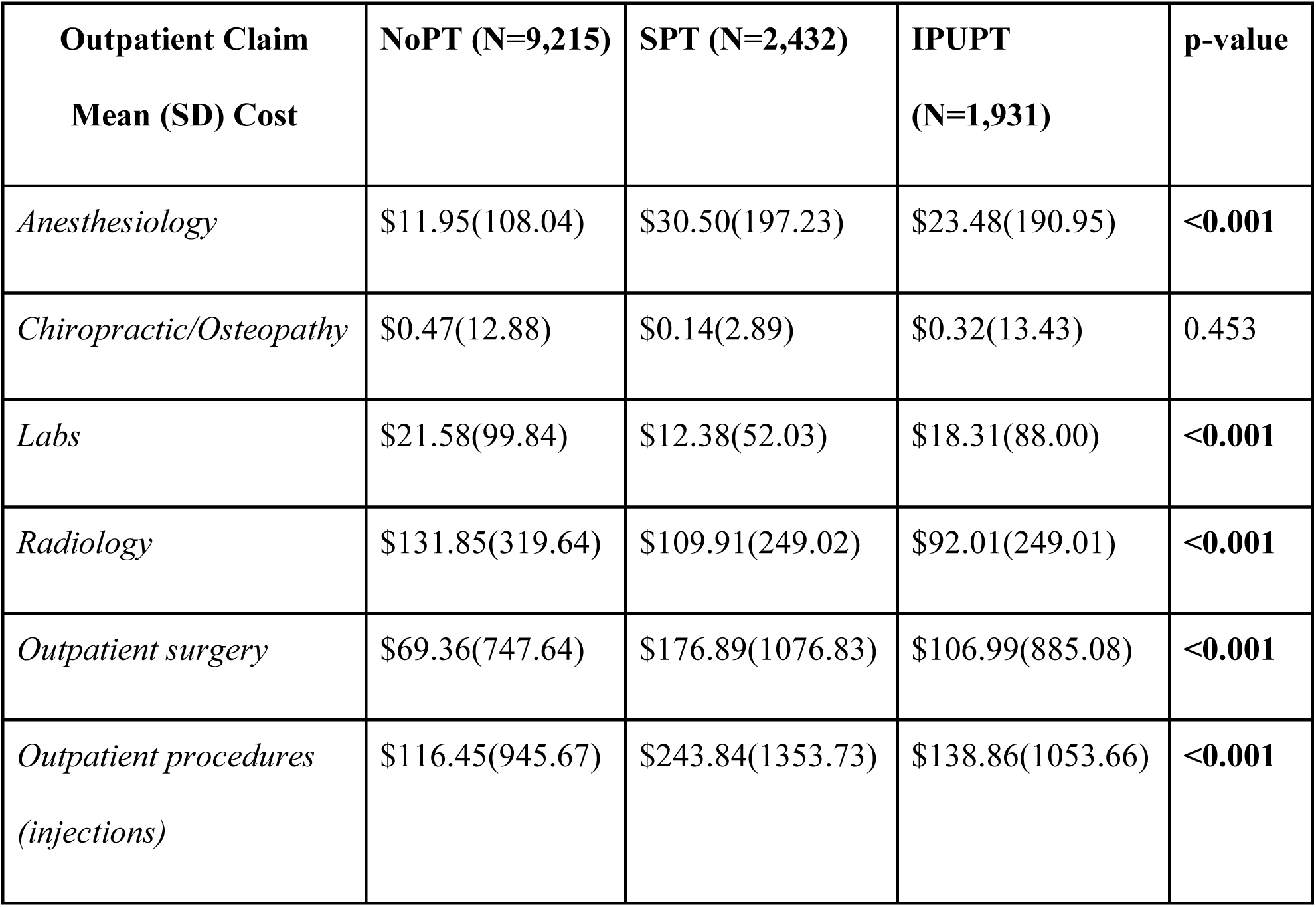
Mean (SD) annual outpatient claim costs per person according to claim category stratified by group. Bold p-values indicate a statistically significant difference in claim costs across groups.

| **Outpatient Claim**<br>**Mean (SD) Cost** | **NoPT (N=9,215)** | **SPT (N=2,432)** | **IPUPT**<br>**(N=1,931)** | **p-value** |
| --- | --- | --- | --- | --- |
| *Anesthesiology* | \$11.95(108.04) | \$30.50(197.23) | \$23.48(190.95) | **<0.001** |
| *Chiropractic/Osteopathy* | \$0.47(12.88) | \$0.14(2.89) | \$0.32(13.43) | 0.453 |
| *Labs* | \$21.58(99.84) | \$12.38(52.03) | \$18.31(88.00) | **<0.001** |
| *Radiology* | \$131.85(319.64) | \$109.91(249.02) | \$92.01(249.01) | **<0.001** |
| *Outpatient surgery* | \$69.36(747.64) | \$176.89(1076.83) | \$106.99(885.08) | **<0.001** |
| *Outpatient procedures*<br>*(injections)* | \$116.45(945.67) | \$243.84(1353.73) | \$138.86(1053.66) | **<0.001** |

## Discussion

This study demonstrated that an IPUPT model was associated with a twofold lower incidence of spine-related inpatient claims when compared to SPT, and was comparable to the NoPT group. Similarly, for outpatient care utilization, the IPUPT group had half of the number of patients utilizing outpatient care compared to the NoPT group, and the SPT group had the greatest number of claims/person per year. Outpatient utilization was primarily driven by radiology costs, followed by laboratory services which were both highest in the NoPT group. Total cost of care per year was over $20 million with the greatest average cost per person in the SPT group at $2,243.66 per person/year. The IPUPT group had the lowest cost of care per person/year at $1,259.88. Outpatient costs represented more than 60% of the total cost of care across the duration of the study period. Membership in the IPUPT group resulted in a total cost saving of approximately $878 per person/year compared to the SPT group, and $16 per person/year compared to the NoPT group, suggesting that not only is the utilization of an interdisciplinary program associated with reduced inpatient and outpatient service utilization, it also reduces overall cost of care over the duration of a 1 year period.

These data are consistent with some prior literature suggesting that initiating physical therapy as a first point of care can reduce costs^5,9^. In this literature, these cost reductions were primarily attributed to reductions in outpatient procedures such as injections and radiology in individuals with acute LBP, which is similar to our findings on the primary drivers of outpatient cost. Another recent study demonstrated small but significant cost savings with clinically significant improvements in disability over a 1-year duration in individuals with acute and subacute back or neck pain who underwent a biopsychosocially informed multidisciplinary intervention as compared to a posturally focused physical therapy intervention or standard conservative care^17^. A recent economic evaluation prepared for the Alliance for Physical Therapy Quality and Innovation reported an overall cost savings of $4,160 with utilization of physical therapy in individuals with acute low back pain as compared to usual care^22^.

Interestingly, although the majority of cost savings with physical therapy have been demonstrated to be in the setting of acute low back pain, the clinical practice guidelines most heavily emphasize physical therapy as a modality to be used for individuals with chronic spine pain, and often omit physical therapy as a recommendation in the acute phase due to the thought that it is primarily self-limiting in the first 90 days^10^. Although our dataset did not include information on chronicity across groups, it is possible that the NoPT group may be biased toward individuals with more acute symptoms, who were not deemed to require a physical therapy referral.

In contrast to the aforementioned evidence supporting cost-effectiveness of physical therapy, two small clinical trials demonstrated no difference in cost-effectiveness of physical therapy as compared to general practitioner’s care alone in individuals with sciatica^23^, and individuals with neck pain^24^. Other trials have found either no difference, or higher total costs in protocols with multidisciplinary protocols, including various combinations of exercise therapy, education, graded activity and behavioral principles as compared to physical therapy in individuals with chronic low back pain^25,26^. However, although more rigorously controlled randomized controlled trials may be useful in evaluating the value of clinical improvements across groups, it may not be representative of actual practice on a larger scale, where individuals often self-select or are directed into care categories by a prescribing primary care provider, and variability in provider and region plays a role in care pathways and cost patterns.

### Limitations

There are several limitations to this study. First, as this was a pragmatic analysis, we did not have demographic or comorbidity data by which to account for potential differences in baseline patient health across groups. It may be likely that individuals in the NoPT group are otherwise healthier and with less comorbidities than those in the physical therapy groups, potentially overestimating equivalent treatment costs for individuals who would otherwise be similarly healthy in the intervention groups. This concept is supported by prior literature demonstrating that individuals with multiple comorbidities are more likely to participate in physical therapy^27^, as well as by the lower non-spine related inpatient costs in the NoPT groups compared to the PT groups. A second limitation is that data on overall quality of life and disability levels for these cohorts was not available, making assessments of clinical value not feasible. Future large scale pragmatic studies including demographic and clinical outcomes data are necessary to understand the value of interdisciplinary care models in physical therapy practice.

## Conclusions

An integrated multidisciplinary rehabilitation model was associated with an 86% lower rates of inpatient spine surgery, a 49% lower cost for spine-related inpatient claims, as well as lower outpatient service utilization compared to SPT and NoPT in individuals with spine pain. The resulting outpatient savings are primarily associated with reductions in radiology and outpatient procedural claims. A significant number of individuals with back pain are receiving spinal surgery without first undergoing physical therapy. Further research is needed to evaluate the impact of an interdisciplinary model on longer term costs and associations with clinical outcomes.

## Data Availability

All data produced in the present study are available upon reasonable request to the authors

## ACKNOWLEDGEMENTS

This study was funded in part by the Foundation for Physical Therapy Research Magistro Family Foundation Grant and NIH R01HD100446. One author discloses a role as Chief Medical Officer of a medical fitness clinic. Due to the de-identified nature of the data utilized in this study, it was determined to be exempt from IRB approval.

## ABBREVIATIONS

LBP: Low Back Pain
NP: Neck pain
IPU: Integrated practice unit
NoPT: no physical therapy group
IPUPT: group receiving physical therapy within an integrated practice unit
SPT: group receiving standard physical therapy
ICD-10: International classification of diseases-10^th^ revision
MSK: musculoskeletal
DRG: diagnosis related group
RVU: relative value unit
CMS: Center for Medicare & Medicaid Services
IPPS: Inpatient Prospective Payment System
CPT: Current procedural terminology

